# Exploring the Relationship Between Acute Respiratory Illnesses, blood inflammatory biomarkers, and Acute Cardiac Events through a cross-sectional study

**DOI:** 10.64898/2026.05.15.26353350

**Authors:** Mohammad Abdul Aleem, C. Raina Macintyre, Bayzidur Rahman, Mohammed Ziaur Rahman, Mustafizur Rahman, A. K. M. Monwarul Islam, Probir Kumar Ghosh, Zubair Akhtar, Fahmida Chowdhury, Firdausi Qadri, Abrar Ahmad Chughtai

**Affiliations:** School of Population Health, UNSW Medicine, the University of New South Wales, Sydney, New South Wales, Australia; Program for Respiratory Infections, Infectious Diseases Division, International Centre for Diarrhoeal Disease Research, Bangladesh (icddr,b), Dhaka, Bangladesh; Biosecurity program, the Kirby Institute, the University of New South Wales, Sydney, New South Wales, Australia; Respiratory and enteric infections, Infectious Diseases Division, International Centre for Diarrhoeal Disease and Research, Bangladesh (icddr,b), Dhaka, Bangladesh; Department of Cardiology, National Institute of Cardiovascular Diseases (NICVD), Dhaka, Bangladesh

**Keywords:** influenza, acute respiratory infection, acute cardiac events, inflammation, biomarkers

## Abstract

**Introduction:** Recent respiratory illness, especially influenza, may trigger acute cardiac events via elevated inflammatory mediators. During the 2018 influenza season in Bangladesh, this study examined whether recent acute clinical respiratory illness (CRI) or laboratory-confirmed influenza was associated with elevated hs-CRP and IL-6, linked to acute cardiac events.

**Methods:** A total of 139 participants aged ≥40 were recruited from a Dhaka cardiac hospital: 70 with acute myocardial infarction (AMI), 30 with other acute cardiac events, and 39 healthy individuals. CRI was defined as fever with cough and/or respiratory symptoms within seven days. Respiratory swabs were tested for influenza, and blood was analyzed for hs-CRP and IL-6.

**Results:** Median hs-CRP and IL-6 were higher in participants with CRI or influenza but not significantly. Cardiac patients had elevated hs-CRP (9.98 mg/L in other cardiac; 4.86 mg/L in AMI vs. 1.73 mg/L in healthy) and IL-6 (0.1 pg/mL in other cardiac; 0.145 pg/mL in AMI vs. 0.08 pg/mL in healthy) (p<0.001). CRI was not significantly associated with elevated hs-CRP or IL-6, though influenza in healthy participants was linked to higher IL-6. Cardiac patients had a higher risk of hs-CRP ≥3 mg/L and elevated IL-6.

**Conclusion:** Cardiac patients showed significantly increased inflammatory markers, but CRI was not clearly linked to inflammation. Further research should assess biomarker utility for early cardiac risk.

**Highlights:**

- The study explores inflammatory biomarkers linking recent respiratory illness and acute cardiac events.
- Cardiac patients showed elevated hs-CRP and IL-6, confirming inflammation–cardiac risk association.
- Association between prior influenza infection and high IL-6 was significant among healthy individuals as opposed to the cardiac group, suggesting effect modification.
- hs-CRP and IL-6 may indicate underlying cardiac pathology and potential early cardiac risk markers.

## ARTICLE SUMMARY

### Key Strengths

- First study in a low-income setting (Bangladesh) examining links between inflammatory biomarkers, respiratory illness, and cardiac events
- Inclusion of diverse study groups (cardiac patients and healthy individuals)
- Participants stratified by recent respiratory illness and laboratory-confirmed influenza status
- Simultaneous assessment of respiratory infections and cardiac conditions within the same population
- Focus on well-established inflammatory biomarkers (hs-CRP and IL-6) relevant to cardiac risk
- Provides novel insights into the relationship between inflammation, respiratory infection, and cardiac events

### Limitations

- Cross-sectional design limits ability to establish causality
- Small sample size, reducing generalizability
- Incomplete laboratory confirmation of respiratory infections in all participants
- Potential misclassification bias due to subjective and non-specific CRI definitions
- Lack of cardiac screening (e.g., ECG, ETT) in healthy participants → possible undetected subclinical disease
- Single time-point (baseline) cytokine measurement, not capturing peak levels
- Absence of a standardized sampling window, leading to variability in cytokine levels
- Dynamic nature of cytokines may cause measurement inaccuracies and false interpretation
- Difficulty distinguishing chronic vs acute inflammatory elevations

### What is already known on this topic?

- In animal models, acute respiratory infections are associated with elevated levels of pro-inflammatory cellular and humoral mediators in both lung and cardiac tissues.
- Epidemiological studies show that increased blood inflammatory markers are linked to adverse respiratory and cardiac outcomes, including ICU admission and death.
- There is a lack of epidemiological evidence on how recent mild to moderate acute respiratory illnesses affect blood cytokine levels compared to individuals without recent illness.
- It is unclear whether changes in cytokine levels following recent respiratory infections increase the risk of imminent acute cardiac events.

### How this study might affect research, practice, or policy?

- This exploratory study is among the first to examine blood inflammatory cytokine patterns within a unified analytical framework, assessing their relationship with both recent acute respiratory illness and acute cardiac events.
- The study observed a slight, though not statistically significant, increase in blood inflammatory mediators following recent acute respiratory illness, indicating the need for further investigation.
- A significant association between recent acute respiratory illness and elevated blood inflammatory mediators was found among otherwise healthy individuals.
- Blood inflammatory mediator levels were significantly higher in patients with acute cardiac conditions compared to healthy individuals.
- These findings highlight the need for further research to determine whether recent acute respiratory illness elevates cytokine levels and whether such increases contribute to the risk of imminent acute cardiac events.
- This study provides preliminary data to inform future research on the potential role of blood inflammatory cytokine profiles in assessing the risk of imminent acute cardiac events, particularly among high-risk populations.

## 1. INTRODUCTION

Adverse cardiac events, including acute myocardial infarction (AMI), are leading contributors to global morbidity and mortality. Recent evidence indicates inflammatory processes, with cytokines playing a key role, significantly contribute to the pathophysiology of atherosclerosis, AMI, and other acute cardiac events. Atherosclerosis, a chronic inflammatory condition, progresses silently, with triggers like infections or exertion causing plaque rupture and acute events. Inflammation can also contribute to other cardiac pathologies like myo- or peri-carditis, arrhythmias, and heart failure by disrupting the heart’s electrical system or damaging heart muscle (1–5).

Chronic heart conditions, when exposed to infectious agents like influenza and RSV, may abruptly worsen through inflammatory responses, potentially leading to acute cardiac events. Highlighting their role in cardiovascular complications, as evidenced by their presence in atherosclerotic plaques, these viruses may accelerate the progression and destabilization of atherosclerosis (6–8). Indirect evidence of increased winter cardiovascular morbidity and mortality due to acute respiratory infections (ARI) like influenza suggests a seasonal spike in AMI risk and deaths. Observational and animal studies further reveal a direct link between recent respiratory illnesses, influenza, systemic and arterial inflammation, and acute cardiovascular events (9–14). In addition to ischaemic heart conditions, viral ARIs can cause other cardiac events like myocarditis and arrhythmias through inflammation. However, genetic variants regulating inflammation may increase inherent susceptibility to such events after a viral ARI, emphasizing the importance of personalized medicine and genetic testing for acute cardiac event prevention (15–18).

Inflammatory mediators like CRP, IL-6, TNF-alpha, IL-1, and MMPs drive AMI onset by promoting inflammation, plaque rupture, and cardiovascular damage (19). Viral ARIs, such as those due to influenza, can elevate these cytokines, exacerbating tissue damage, myocarditis, arrhythmias, and heart failure (20–22)(12, 23). Conversely, influenza vaccination has been shown to reduce cardiovascular events. A 2021 study by Rony et al. demonstrated that the influenza vaccine reduces proinflammatory mediators like IL-6, IL-8, and TNF while increasing anti-inflammatory IL-10 post-coronary bypass. These findings, supported by mouse models, suggest the vaccine promotes a stable anti-inflammatory response, further highlighting the potential of inflammatory mediators in cardiac prevention (24–26).

This study aimed to explore whether inflammatory markers can serve as early indicators of acute cardiac events following recent ARI or influenza. The specific objectives of this study were to investigate the relationships of recent acute respiratory illnesses and acute cardiac events, including AMI, with blood inflammatory biomarkers. Additionally, the study evaluated how respiratory illnesses or influenza differentially impact inflammatory biomarker levels in cardiac patients compared to healthy individuals.

## 2. MATERIALS AND METHODS

Participants for the current analysis were randomly selected from a prior case-control study conducted at the National Institute of Cardiovascular Diseases (NICVD) in Dhaka, Bangladesh, during the 2018 influenza season (May 1 to October 31, 2018) (27). The description of annual influenza seasons for Bangladesh based on national surveillance data is discussed in detail by Zaman et al. (28).

### 2.1. Study design and study population

This cross-sectional study (Fig 1 & Fig 2) was conducted among three distinct participant groups: i). Patients hospitalized with AMI ii). Patients admitted with acute cardiac events other than AMI or ischemic heart diseases and iii). Apparently healthy individuals.

**Figure 1.**
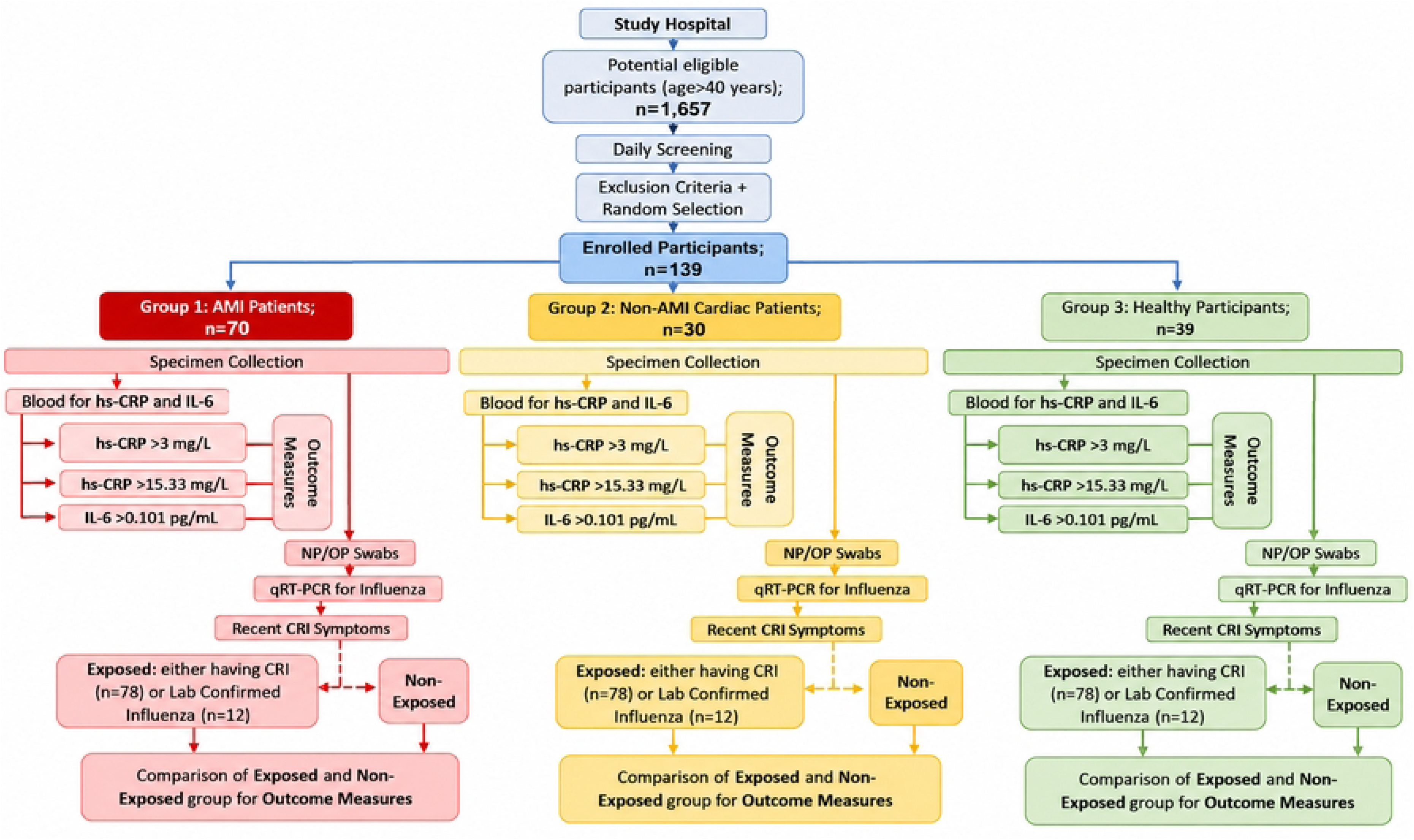
Patient flow diagram

**Figure 2.**
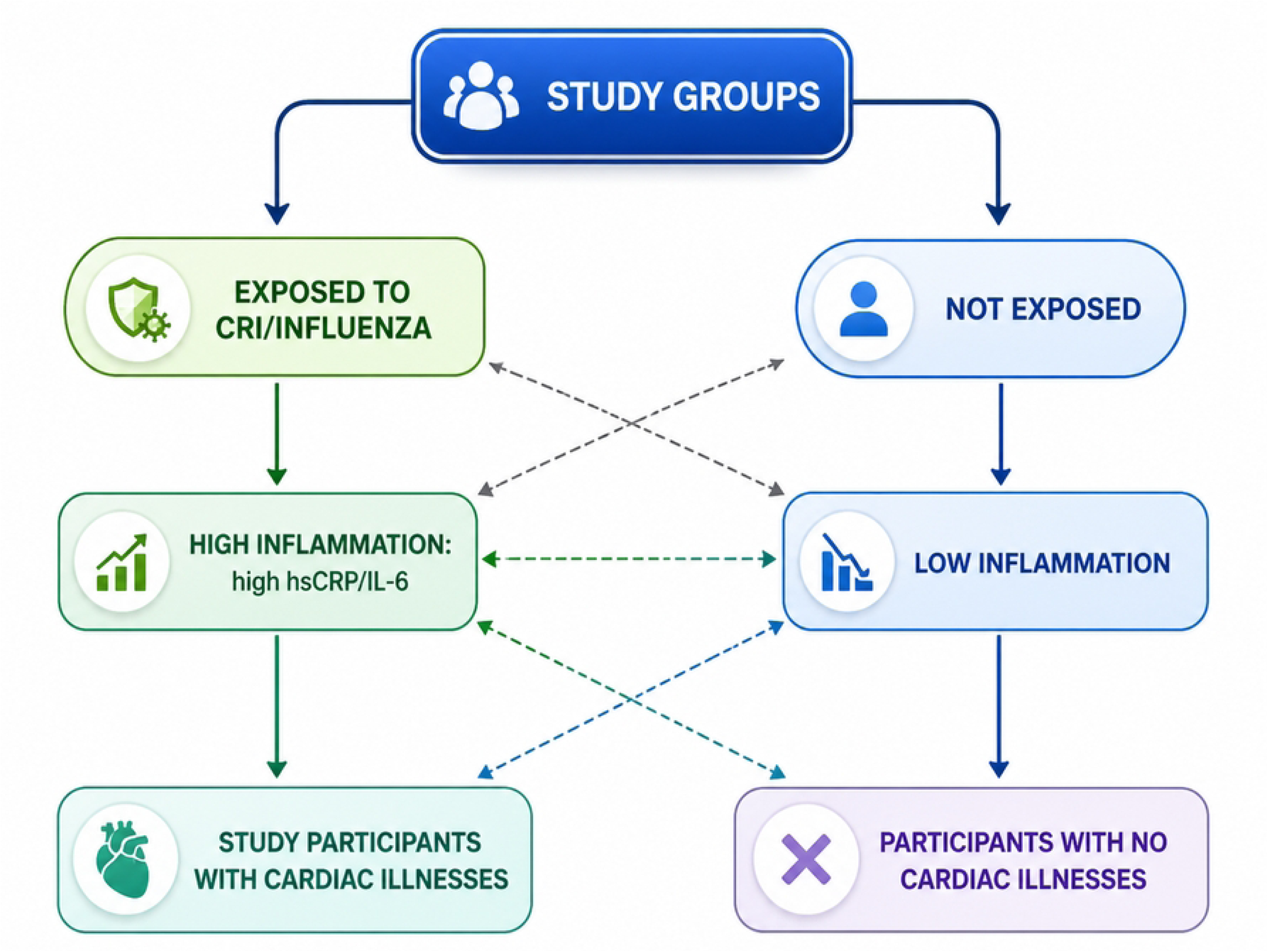
Study design conceptual diagram: Relationships between respiratory exposure, high blood levels of inflammatory biomarkers and cardiac illnesses

### 2.2. Enrolment of study participants and exclusion criteria

The details have been described elsewhere (27). The NICVD study team recruited participants aged 40+ during the 2018 influenza season. A physician conducted daily inpatient rounds, screening for acute myocardial infarction (AMI), other cardiac conditions, and healthy controls. AMI patients were identified through symptoms, ECG, cardiac biomarkers, or coronary interventions, with diagnoses confirmed by cardiologists. Non-AMI cardiac patients included those with conditions like valvular heart disease, hypertension, or cardiomyopathy, verified via medical history and diagnostic tests. Healthy controls were recruited from patient visitors, though subclinical conditions might not have been excluded. Exclusion criteria applied to individuals with chronic liver/renal issues, malignancies, autoimmune/inflammatory diseases, recent infections/vaccinations, NSAID use, or inability to consent.

### 2.3. Data collection

After obtaining consent, study staff reviewed medical records and conducted structured interviews to gather sociodemographic data, lifestyle information, co-morbid illnesses, and recent respiratory symptoms like fever, cough, sore throat, runny nose, and difficulty breathing. Physical measurements, including clinical examinations and anthropometrics, assessed participants’ height, weight, BMI, and overall health.

### 2.4. Specimen collection and laboratory testing

Trained staff collected nasopharyngeal (NP) and oropharyngeal (OP) swabs from all participants after enrolment. Specifically, for patients, swabs were collected within 72 hours of the onset of the cardiac event to enhance influenza RNA detection before viral shedding ceased. Swabs were transported in Viral Transport Medium (VTM) to the icddr,b Virology Laboratory and stored at −70°C. RNA was extracted, and qRT-PCR was used to detect influenza A and B viruses. Hemagglutinin subtyping identified specific strains, following validated protocols and standard quality control procedures.

Blood samples were collected from all participants upon enrolment, with patient samples taken specifically within 72 hours of the onset of cardiac events. Blood samples were collected in both plain and plasma separator tubes, and stored in iceboxes at a temperature of 2-8°C. A trained Medical Technologist (MT) with prior experience collected the blood samples. Blood samples were collected using a vacutainer and butterfly needle. The participant’s vein was cleaned, and a tourniquet was applied to highlight veins. A butterfly needle was inserted, and blood flowed into the vacutainer. After collection, the tourniquet was released, and pressure applied. Samples were centrifuged, labelled, and sent for analysis. The serum samples were tested for hs-CRP, while the plasma samples were tested for IL-6 using commercially available ELISA kits in the biochemistry and immunology laboratories of icddr,b, respectively. The laboratory analysis of the inflammatory mediators was conducted following the instructions provided by the ELISA kit manufacturers.

### 2.5. Exposure measures

The first primary exposure in this study was recent Clinical Respiratory Illness (CRI), defined as a self-reported history of two or more respiratory symptoms (fever, cough, sore throat, runny nose, or difficulty breathing) within the past seven days (27, 29–31). The second primary exposure was qRT-PCR confirmed influenza A or B identified in the respiratory swabs. The participants were divided into two groups based on their history of recent CRI and influenza test results, with those who had a history of recent CRI or laboratory-confirmed influenza considered as “exposed,” and those who didn’t report recent CRI or influenza were considered as “non-exposed.“

### 2.6. Outcome measures

Outcome variables in the current study were high-risk levelsof hs-CRP and IL-6. For hs-CRP, blood concentrations either above clinical cut-off value of 3 mg/L (32–34) or greater than 4^th^ quartile threshold value (35) of 15.33 mg/L were used to define high-risk levels. For IL-6 blood levels above the median cut-off value (36) of 0.1010 pg/mL were considered high risk. For IL-6 the median cut-off value was used instead of 4^th^ quartile threshold value due to small sample size. Baseline blood concentration of inflammatory mediators were compared between the “exposed” and “non- exposed” groups, with a focus on whether the concentration was above or below cut off value for both groups.

### 2.7. Data analysis

Chi-squared or Fisher’s exact tests were used to compare categorical variables, including sociodemographic, clinical, CRI, and influenza status. Baseline inflammatory mediator levels were examined via histograms and box plots. Due to skewed distributions, medians with IQRs were used instead of means, and comparisons were made using the Mann-Whitney U test for two groups and the Kruskal-Wallis test for more than two groups. Medians were compared between cardiac and healthy participants, as well as those positive or negative for CRI or influenza. Although mean values were also examined, no statistical test was performed on them. Finally, inflammatory mediator levels were categorized into risk groups using previously established cutoff values.

Simple and multivariable log-binomial regression was used to investigate the relationship between recent CRI, influenza status, and high-risk blood inflammatory mediators (IL-6 or hs-CRP) and to estimate crude and adjusted relative risk ratios (RRs and aRRs). The multivariable model adjusted for sociodemographic and clinical covariates, including age, sex, education, smoking status, physical activity, BMI, blood lipid levels, hypertension, diabetes, and participant groups (healthy vs. cardiac, with the cardiac group encompassing all cardiac illnesses, including AMI). Participants were initially included in the model if variables had a p-value <0.25. A backward elimination method, guided by likelihood ratio tests, refined the model by removing non-significant variables until only those with p-values <0.05 or identified as confounders (causing a ≥10% change in association) remained. Multicollinearity was assessed using the Variance Inflation Factor (VIF), with VIF >5 indicating significant multicollinearity, necessitating the removal of correlated variables.

The study also examined whether cardiac conditions modify how CRI or influenza influences high-risk inflammatory biomarkers. To assess effect modification, interaction terms were added between CRI and participant group (cardiac vs. healthy) and between influenza and participant group in multivariable models. A significant interaction (p<0.05) indicated modification by participant group. For associations showing significant interaction, aRRs were estimated separately for the cardiac and healthy groups. All analyses were performed using SAS version 9.4.

### 2.8. Sample size calculation

To assess the association between recent respiratory illness and high blood inflammatory mediators (IL-6 or hs-CRP) in this cross-sectional study, a sample size of at least 118 participants (59 exposed, 59 non-exposed) was calculated using STATA 13, aiming for 80% power and p < 0.05, assuming 5% high-risk in the exposed group and 2.5% in the non-exposed group. The study ultimately included 139 participants: 78 with recent CRI and 61 without, consisting of 70 AMI patients, 30 with non-ischemic cardiac conditions, and 39 healthy individuals.

### 2.9. Ethical approval

Enrollment of the participants started after approval of the study by the icddr,b Institutional Review Board (PR-17039) and UNSW Human Research Ethics Committee (HC 17861). Informed written consent to participate in the study was obtained. Consent was obtained by the study physician using a detailed consent form, ensuring voluntary participation and the right to withdraw. Participants received explanations, could ask questions, and got copies of the signed form. The process prioritized autonomy, protected rights, and maintained confidentiality.

## 3. Results

Across AMI, other cardiac, and healthy participants, education levels were lower in the cardiac groups (p=0.03). Hypertension was most common in AMI patients (45.7%), while diabetes was highest among healthy individuals (23.1%) (p<0.001). Tobacco use, though elevated among AMI patients (75.7%), was not significant. Healthy participants had more family history of CVD (p=0.01). There were no significant differences in BMI, cholesterol, CRI, or influenza status (p>0.22). Influenza positivity was highest among AMI patients (10%) compared to cardiac (6.7%) and healthy participants (7.7%).

Both hs-CRP and IL-6 levels were significantly different between the groups (P < 0.001) (Table 1). The cardiac patient group had the highest median hs-CRP level (9.98 mg/L), while the AMI patient group had the highest median IL-6 level (0.145 pg/mL). AMI and cardiac patients exhibited higher hs-CRP and IL-6 levels compared to healthy participants, who had median levels of 1.73 mg/L for hs-CRP and 0.08 pg/mL for IL-6.

**Table 1.** Sociodemographic and clinical characteristics of participant groups.

| **Sociodemographic and clinical characteristics** | **AMI patients, N=70, n (%)** | **Cardiac patients, N=30, n (%)** | **Healthy participants, N=39, n (%)** | **p-value\*** |
| --- | --- | --- | --- | --- |
| Age |  |  |  |  |
| 40-64 years | 57 (81.4) | 21 (70.0) | 34 (87.2) |  |
| ≥65 years | 13 (18.6) | 9 (30.0) | 5 (12.8) | 0.20 |
| Gender |  |  |  |  |
| Male | 67 (95.7) | 29 (96.7) | 38 (97.4) |  |
| Female | 3 (4.3) | 1 (3.3) | 1 (2.6) | 0.90 |
| Education |  |  |  |  |
| <primary | 35 (50.0) | 18 (60.0) | 11 (28.2) |  |
| primary to HSC | 33 (47.1) | 11 (36.7) | 23 (59.0) |  |
| Completed graduation | 2 (2.9) | 1 (3.3) | 5 (12.8) | 0.03 |
| Past medical history |  |  |  |  |
| No | 29 (41.4) | 7 (23.3) | 20 (51.3) |  |
| Yes | 41 (58.6) | 23 (76.7) | 19 (48.7) | 0.06 |
| Hypertension |  |  |  |  |
| No | 38 (54.3) | 14 (46.7) | 21 (53.8) |  |
| Yes | 32 (45.7) | 9 (30.0) | 12 (30.8) | <0.001 |
| Unknown | 0 (0.0) | 7 (23.3) | 6 (15.4) |  |
| Diabetes |  |  |  |  |
| No | 61 (87.1) | 19 (63.3) | 22 (56.4) |  |
| Yes | 9 (12.9) | 4 (13.3) | 9 (23.1) |  |
| Unknown | 0 (0.0) | 7 (23.3) | 8 (20.5) | <0.001 |
| Tobacco use |  |  |  |  |
| No | 17 (24.3) | 12 (40.0) | 15 (38.5) |  |
| Yes | 53 (75.7) | 18 (60.0) | 24 (61.5) | 0.17 |
| Physical activities |  |  |  |  |
| No | 33 (47.1) | 17 (56.7) | 15 (38.5) |  |
| Yes | 37 (52.9) | 13 (43.3) | 24 (61.5) | 0.32 |
| Family history of CVD |  |  |  |  |
| No | 26 (37.1) | 16 (53.3) | 8 (20.5) |  |
| Yes | 44 (62.9) | 14 (46.7) | 31 (79.5) | 0.01 |
| BMI |  |  |  |  |
| <18.5 | 15 (21.4) | 5 (16.7) | 2 (5.1) |  |
| 18.5-24.9 | 32 (45.7) | 16 (53.3) | 23 (59.0) |  |
| 25-29.9 | 19 (27.1) | 6 (20.0) | 13 (33.3) |  |
| ≥30 | 4 (5.7) | 2 (6.7) | 0 (0.0) | 0.22 |
| High blood cholesterol |  |  |  |  |
| No | 56 (80.0) | 24 (80.0) | 29 (74.4) |  |
| Yes | 14 (20.0) | 6 (20.0) | 10 (25.6) | 0.77 |
| CRI status |  |  |  |  |
| Without CRI | 29 (41.4) | 12 (40.0) | 20 (51.3) |  |
| With CRI | 41 (58.6) | 18 (60.0) | 19 (48.7) | 0.54 |
| Influenza status |  |  |  |  |
| Influenza negative | 63 (90.0) | 28 (93.3) | 36 (92.3) |  |
| Influenza positive | 7 (10.0) | 2 (6.7) | 3 (7.7) | 0.84 |
| Median (IQR) hs-CRP level in mg/L | 4.86 (2.66-15.75) | 9.98 (2.73-29.23) | 1.73 (0.54-2.99) | <0.001** |
| Median IL-6 (IQR) level in pg/mL | 0.145 (0.099-0.258) | 0.1 (0.08-0.190) | 0.08 (0.075-0.087) | <0.001** |

\* Chi-square test or Fisher's exact test performed to compare percentages across independent groups
\*\* Kruskal-Wallis test performed to compare the median levels between the three groups of participants

### 3.1. Blood inflammatory markers among individuals with clinical respiratory illness and influenza

The **Fig 3A & Fig 3B** presents a comparison of hs-CRP and IL-6 levels in relation to CRI and influenza status. Individuals with CRI or influenza had consistently higher median and mean levels of hs-CRP and IL-6 compared to those without CRI and influenza. However, the differences between the median levels were statistically not significant. Median hs-CRP levels were 4.53 mg/L in CRI participants versus 3.25 mg/L without CRI, and 8.12 mg/L in influenza-positive versus 3.52 mg/L negative participants (p=0.17 and 0.24, respectively). Median IL-6 levels showed similar trends (p=0.63 and 0.18). Differences were more pronounced in the upper quartile, suggesting CRI and influenza may lead to elevated and variable inflammatory markers in some individuals.

**Figure 3A.**
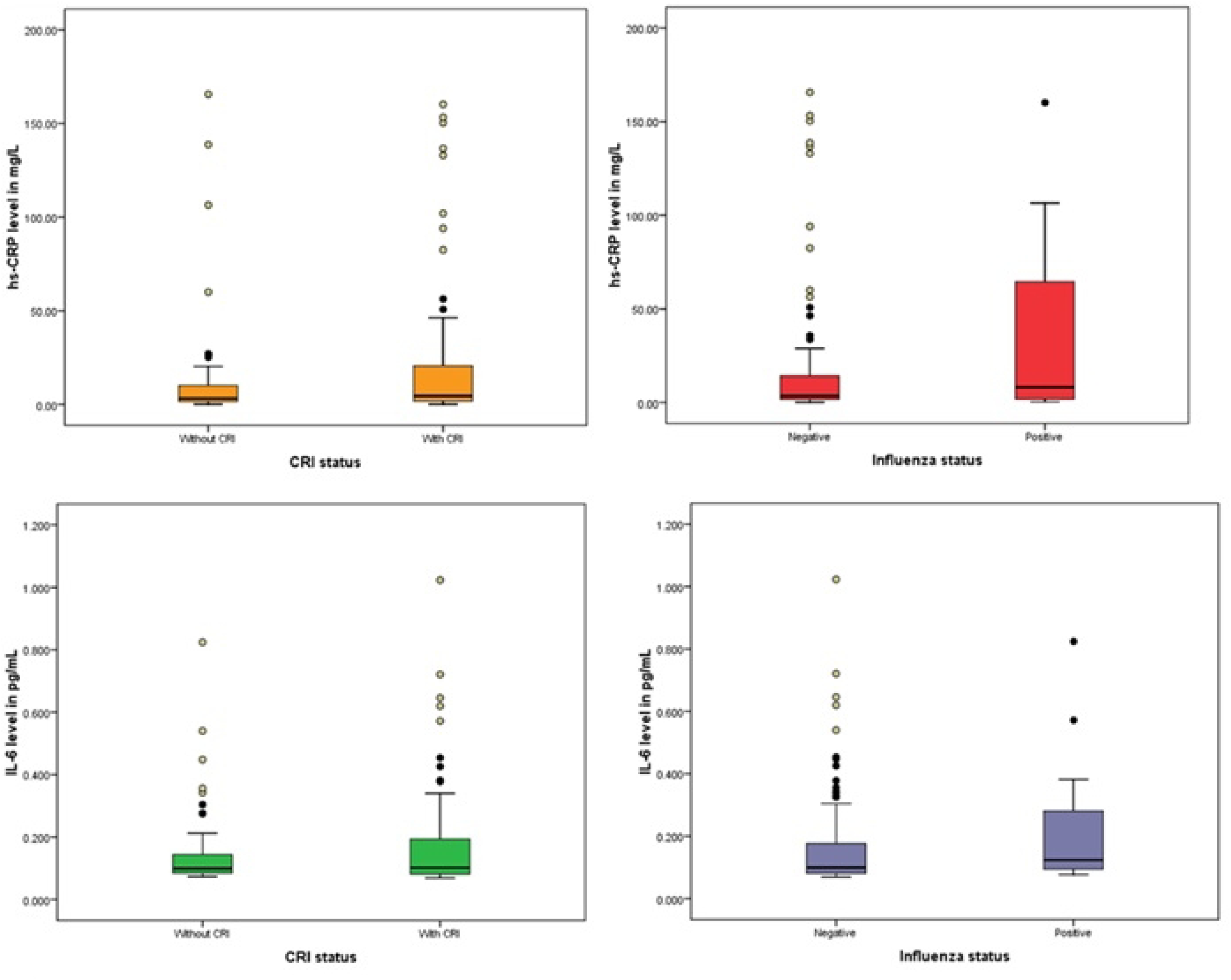
Comparison of median (IQR) levels of hs-CRP (mg/L) and IL-6 (pg/mL) in relation to clinical respiratory illness and influenza status

**Note:**

– Number of participants by CRI and influenza status:

- Without CRI (N=61); With CRI (N=78)
- Influenza negative (N=127), Influenza positive (N=12)
– Mann-Whitney U test performed to determine the statistical significance of the differences in medians
– p-values:

- **hs-CRP**: CRI (p=0.17), Influenza (p=0.24)
- **IL-6**: CRI (p=0.63), Influenza (p=0.18)

**Figure 3B.**
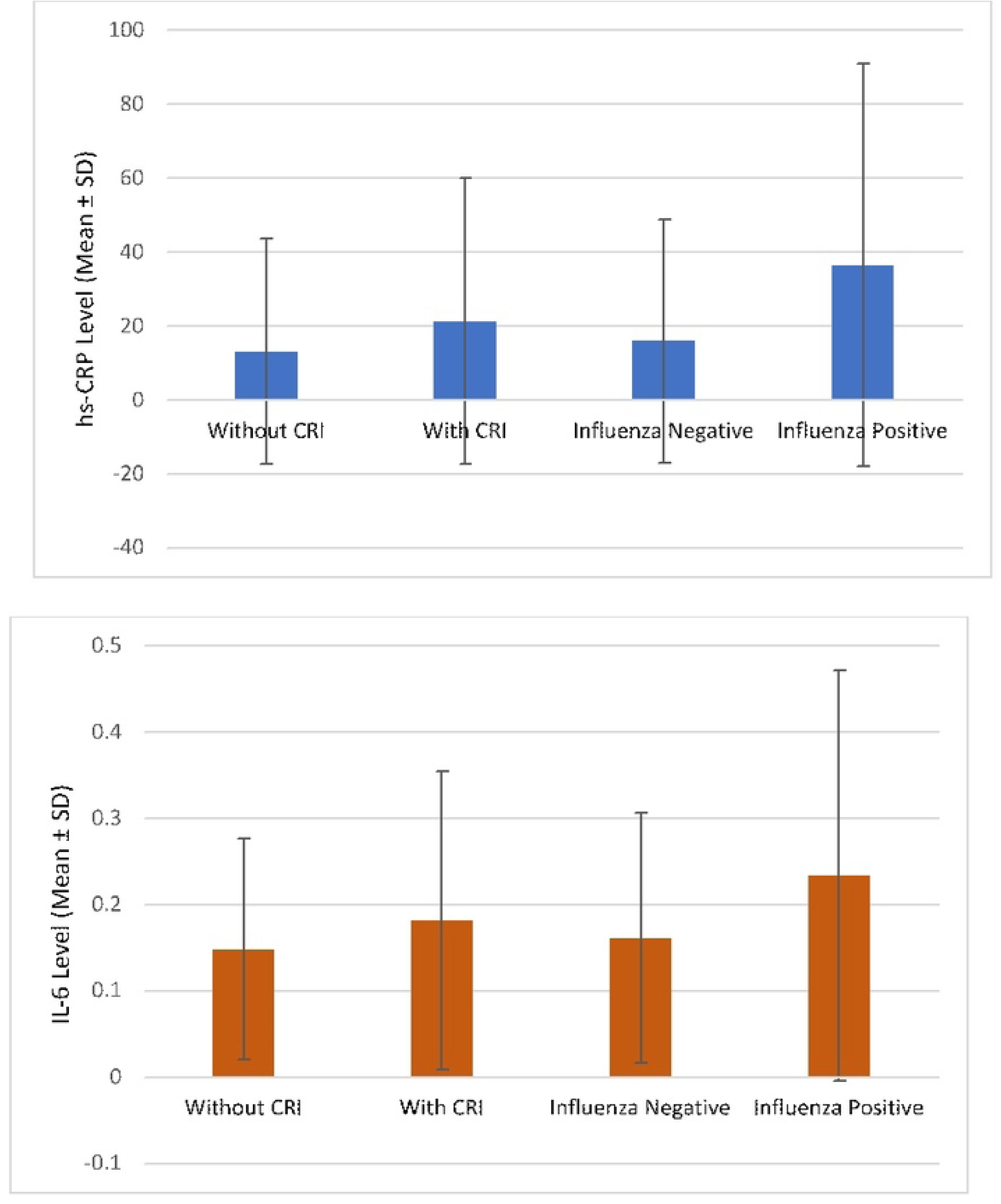
Comparison of mean (SD) levels of hs-CRP (mg/L) and IL-6 (pg/mL) in relation to clinical respiratory illness and influenza status

**Note:**

– No statistical tests for significance was performed
– Error bars represent SDs

### 3.2. Association of recent clinical respiratory illness and influenza with high-risk hs-CRP blood level

There was no significant association between CRI status and high-risk hs-CRP level ≥3 mg/L (aRR: 0.99; 95% CI: 0.82, 1.19) or ≥4th Quartile (aRR: 1.41; 95% CI: 0.75, 2.67) after adjusting for potential confounders **(Table 2)**. However, the multivariable model showed the cardiac group was significantly more likely to have high-risk hs-CRP levels with aRR (95% CI) as 1.54 (1.15, 2.04) and 4.03 (1.27, 12.79) compared to the healthy participants. **Table 3** presents the results of the multivariable model examining the relationship of influenza status with high-risk hs-CRP level ≥3 mg/L and ≥4th Quartile. None of the associations were significant. The aRRs (95% CIs) were 1.17 (0.89–1.54) and 1.88 (0.85–4.17) respectively. However, the model showed that the cardiac patients’ group had a significantly higher risk of having high hs-CRP levels compared to the healthy participants aRRs (95% CIs) were 1.55 (1.17-2.05) and 3.86 (1.24–11.97).

**Table 2.**
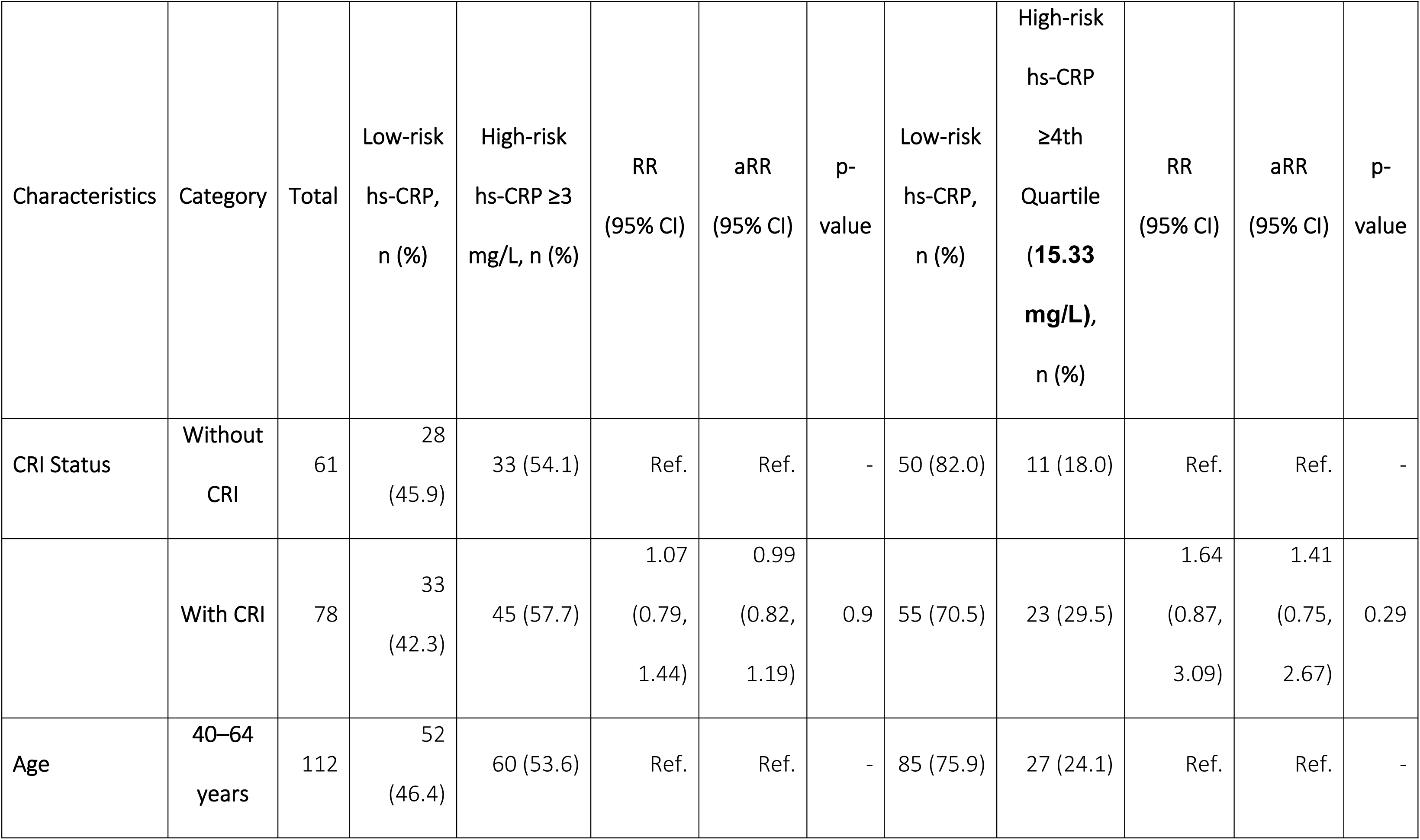
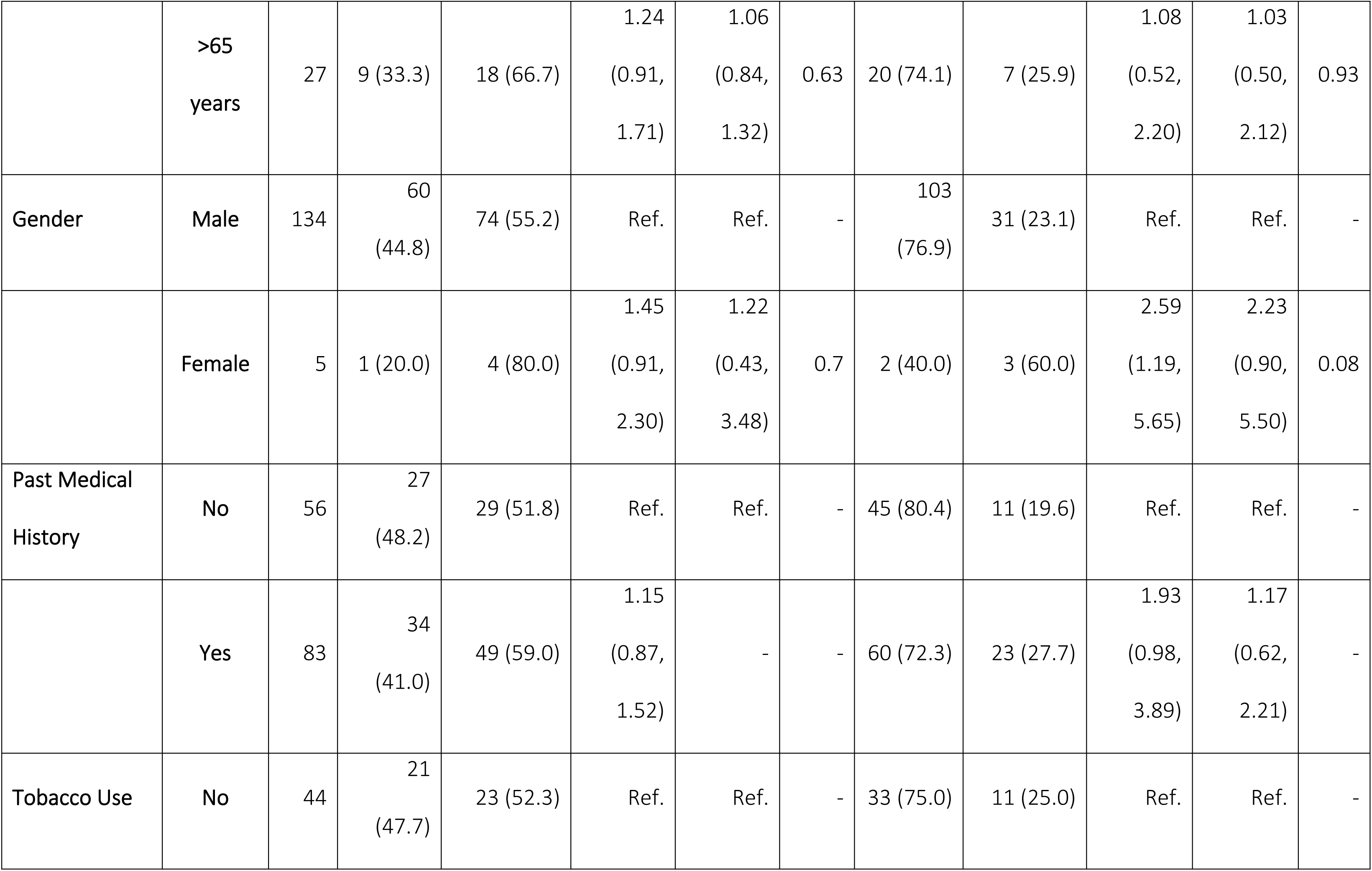
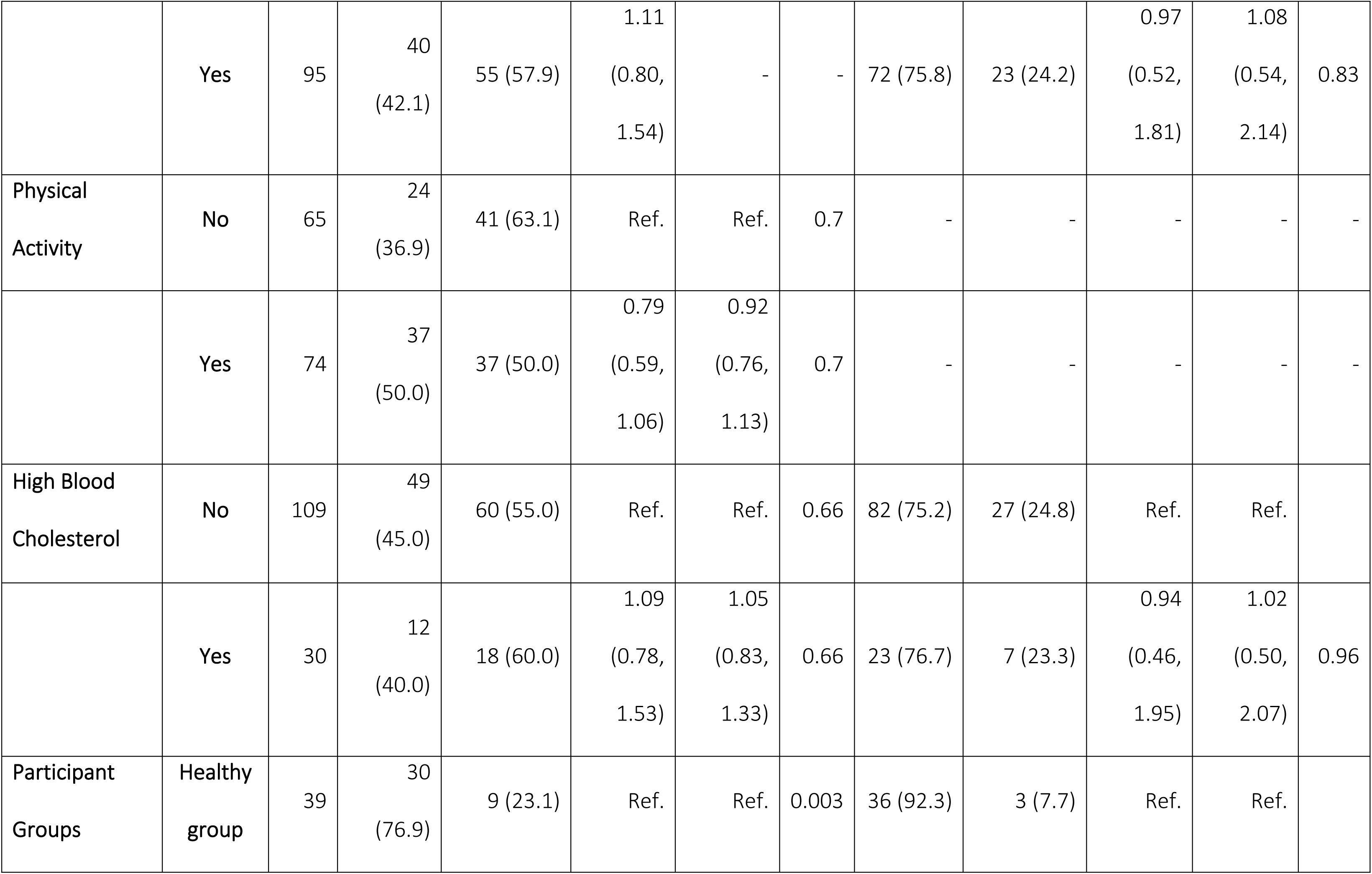
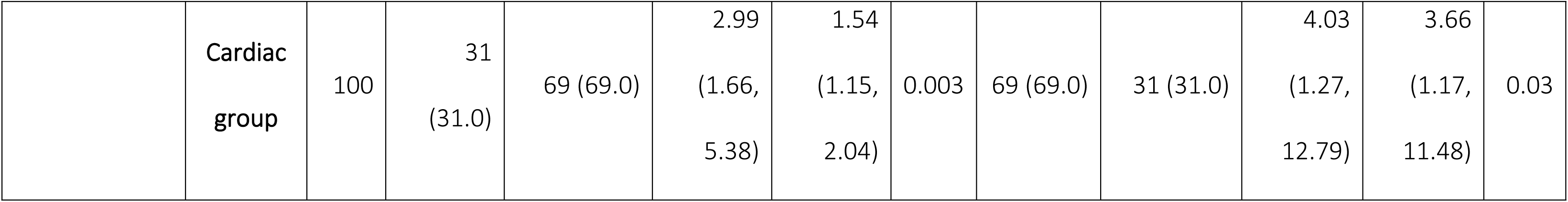
Association of recent clinical respiratory illness status with high-risk hs-CRP levels.

| Characteristics | Category | Total | Low-risk<br>hs-CRP,<br>n (%) | High-risk<br>hs-CRP ≥3<br>mg/L, n (%) | RR<br>(95% CI) | aRR<br>(95% CI) | p-value | Low-risk<br>hs-CRP,<br>n (%) | High-risk<br>hs-CRP<br>≥4th<br>Quartile<br>(15.33<br>mg/L),<br>n (%) | RR<br>(95% CI) | aRR<br>(95% CI) | p-value |
| --- | --- | --- | --- | --- | --- | --- | --- | --- | --- | --- | --- | --- |
| CRI Status | Without<br>CRI | 61 | 28<br>(45.9) | 33 (54.1) | Ref. | Ref. | - | 50 (82.0) | 11 (18.0) | Ref. | Ref. | - |
|  | With CRI | 78 | 33<br>(42.3) | 45 (57.7) | 1.07<br>(0.79,<br>1.44) | 0.99<br>(0.82,<br>1.19) | 0.9 | 55 (70.5) | 23 (29.5) | 1.64<br>(0.87,<br>3.09) | 1.41<br>(0.75,<br>2.67) | 0.29 |
| Age | 40–64<br>years | 112 | 52<br>(46.4) | 60 (53.6) | Ref. | Ref. | - | 85 (75.9) | 27 (24.1) | Ref. | Ref. | - |
|  | >65<br>years | 27 | 9 (33.3) | 18 (66.7) | 1.24<br>(0.91,<br>1.71) | 1.06<br>(0.84,<br>1.32) | 0.63 | 20 (74.1) | 7 (25.9) | 1.08<br>(0.52,<br>2.20) | 1.03<br>(0.50,<br>2.12) | 0.93 |
| Gender | Male | 134 | 60<br>(44.8) | 74 (55.2) | Ref. | Ref. | - | 103<br>(76.9) | 31 (23.1) | Ref. | Ref. | - |
|  | Female | 5 | 1 (20.0) | 4 (80.0) | 1.45<br>(0.91,<br>2.30) | 1.22<br>(0.43,<br>3.48) | 0.7 | 2 (40.0) | 3 (60.0) | 2.59<br>(1.19,<br>5.65) | 2.23<br>(0.90,<br>5.50) | 0.08 |
| Past Medical<br>History | No | 56 | 27<br>(48.2) | 29 (51.8) | Ref. | Ref. | - | 45 (80.4) | 11 (19.6) | Ref. | Ref. | - |
|  | Yes | 83 | 34<br>(41.0) | 49 (59.0) | 1.15<br>(0.87,<br>1.52) | - | - | 60 (72.3) | 23 (27.7) | 1.93<br>(0.98,<br>3.89) | 1.17<br>(0.62,<br>2.21) | - |
| Tobacco Use | No | 44 | 21<br>(47.7) | 23 (52.3) | Ref. | Ref. | - | 33 (75.0) | 11 (25.0) | Ref. | Ref. | - |
|  | Yes | 95 | 40<br>(42.1) | 55 (57.9) | 1.11<br>(0.80,<br>1.54) | - | - | 72 (75.8) | 23 (24.2) | 0.97<br>(0.52,<br>1.81) | 1.08<br>(0.54,<br>2.14) | 0.83 |
| Physical Activity | No | 65 | 24<br>(36.9) | 41 (63.1) | Ref. | Ref. | 0.7 | - | - | - | - | - |
|  | Yes | 74 | 37<br>(50.0) | 37 (50.0) | 0.79<br>(0.59,<br>1.06) | 0.92<br>(0.76,<br>1.13) | 0.7 | - | - | - | - | - |
| High Blood Cholesterol | No | 109 | 49<br>(45.0) | 60 (55.0) | Ref. | Ref. | 0.66 | 82 (75.2) | 27 (24.8) | Ref. | Ref. |  |
|  | Yes | 30 | 12<br>(40.0) | 18 (60.0) | 1.09<br>(0.78,<br>1.53) | 1.05<br>(0.83,<br>1.33) | 0.66 | 23 (76.7) | 7 (23.3) | 0.94<br>(0.46,<br>1.95) | 1.02<br>(0.50,<br>2.07) | 0.96 |
| Participant Groups | Healthy group | 39 | 30<br>(76.9) | 9 (23.1) | Ref. | Ref. | 0.003 | 36 (92.3) | 3 (7.7) | Ref. | Ref. |  |
|  | Cardiac<br>group | 100 | 31<br>(31.0) | 69 (69.0) | 2.99<br>(1.66,<br>5.38) | 1.54<br>(1.15,<br>2.04) | 0.003 | 69 (69.0) | 31 (31.0) | 4.03<br>(1.27,<br>12.79) | 3.66<br>(1.17,<br>11.48) | 0.03 |

**Table 3.** Association of influenza status with high-risk hs-CRP levels.

| Characteristics | Category | Total | Low-risk<br>hs-CRP,<br>n (%) | High-risk<br>hs-CRP<br>≥3 mg/L,<br>n (%) | RR<br>(95% CI) | aRR<br>(95% CI) | p-value | Low-risk<br>hs-CRP,<br>n (%) | High-risk<br>hs-CRP<br>≥4th<br>Quartile<br>(15.33<br>mg/L),<br>n (%) | RR<br>(95% CI) | aRR<br>(95% CI) | p-value |
| --- | --- | --- | --- | --- | --- | --- | --- | --- | --- | --- | --- | --- |
| Influenza<br>Status | Influenza<br>Negative | 127 | 58 (45.7) | 69 (54.3) | Ref. | Ref. | — | 98<br>(77.2) | 29 (22.8) | Ref. | Ref. | — |
|  | Influenza<br>Positive | 12 | 3 (25.0) | 9 (75.0) | 1.38<br>(0.96–<br>1.99) | 1.17<br>(0.89–<br>1.54) | 0.24 | 7 (58.3) | 5 (41.7) | 1.82<br>(0.87–<br>3.83) | 1.88<br>(0.85–<br>4.17) | 0.12 |
| Age | 40–64<br>years | 112 | 52 (46.4) | 60 (53.6) | Ref. | Ref. | — | 85<br>(75.9) | 27 (24.1) | Ref. | Ref. | — |
|  | >65<br>years | 27 | 9 (33.3) | 18 (66.7) | 1.24<br>(0.91–<br>1.71) | 1.06<br>(0.84–<br>1.34) | 0.62 | 20<br>(74.1) | 7 (25.9) | 1.08<br>(0.52–<br>2.20) | 1.08<br>(0.51–<br>2.28) | 0.84 |
| Gender | Male | 134 | 60 (44.8) | 74 (55.2) | Ref. | Ref. | — | 103<br>(76.9) | 31 (23.1) | Ref. | Ref. | — |
|  | Female | 5 | 1 (20.0) | 4 (80.0) | 1.45<br>(0.91–<br>2.30) | 1.23<br>(0.42–<br>3.53) | 0.71 | 2 (40.0) | 3 (60.0) | 2.59<br>(1.19–<br>5.65) | 2.60<br>(1.04–<br>6.55) | 0.04 |
| Past Medical<br>History | No | 56 | 27 (48.2) | 29 (51.8) | Ref. | — | — | 45<br>(80.4) | 11 (19.6) | Ref. | Ref. | — |
|  | Yes | 83 | 34 (41.0) | 49 (59.0) | 1.15<br>(0.87–<br>1.52) | — | — | 60<br>(72.3) | 23 (27.7) | 1.93<br>(0.98–<br>3.89) | 1.15<br>(0.61–<br>2.17) | 0.66 |
| Tobacco Use | No | 44 | 21 (47.7) | 23 (52.3) | Ref. | — | — | 33<br>(75.0) | 11 (25.0) | Ref. | Ref. | — |
|  | Yes | 95 | 40 (42.1) | 55 (57.9) | 1.11<br>(0.80–<br>1.54) | — | — | 72<br>(75.8) | 23 (24.2) | 0.97<br>(0.52–<br>1.81) | 0.96<br>(0.47–<br>1.96) | 0.92 |
| Physical<br>Activities | No | 65 | 24 (36.9) | 41 (63.1) | Ref. | Ref. | 0.46 | 50<br>(76.9) | 15 (23.1) | Ref. | — | — |
|  | Yes | 74 | 37 (50.0) | 37 (50.0) | 0.79<br>(0.59–<br>1.06) | 0.93<br>(0.77–<br>1.13) | 0.46 | 55<br>(74.3) | 19 (25.7) | 1.11<br>(0.62–<br>2.01) | — | — |
| High Blood<br>Cholesterol | No | 109 | 49 (45.0) | 60 (55.0) | Ref. | Ref. | 0.6 | 82<br>(75.2) | 27 (24.8) | Ref. | Ref. | 0.77 |
|  | Yes | 30 | 12 (40.0) | 18 (60.0) | 1.09<br>(0.78–<br>1.53) | 1.06<br>(0.85–<br>1.32) | 0.6 | 23<br>(76.7) | 7 (23.3) | 0.94<br>(0.46–<br>1.95) | 1.12<br>(0.54–<br>2.32) | 0.77 |
| Participant<br>Groups | Healthy<br>group | 39 | 30 (76.9) | 9 (23.1) | Ref. | Ref. | 0.002 | 36<br>(92.3) | 3 (7.7) | Ref. | Ref. | 0.02 |
|  | Cardiac<br>group | 100 | 31 (31.0) | 69 (69.0) | 2.99<br>(1.66–<br>5.38) | 1.55<br>(1.17–<br>2.05) | 0.002 | 69<br>(69.0) | 31 (31.0) | 4.03<br>(1.27–<br>12.79) | 3.86<br>(1.24–<br>11.97) | 0.02 |

### 3.3. Association of clinical respiratory illness and influenza status with high-risk IL-6 blood level

There was no significant association between CRI status and high IL-6 levels (aRR: 0.97, 95% CI: 0.72-1.29) (Table 4). However, a strong association was found between the cardiac group and high IL-6 levels (aRR: 4.84, 95% CI: 2.11-11.16). Using multivariable models with interaction terms, we found that only the association between influenza and IL-6 levels was significantly modified by participant groups (p = 0.04).Only the aRR for this association within the healthy participants’ group was significant (aRR: 8.18, 95% CI: 1.34- 49.82).

**Table 4.** Association of recent clinical respiratory illness status and influenza status with high-risk IL-6 Level.

| Characteristics | Category | Total | Low<br>IL-6,<br>n (%) | High IL-6,<br>≥<br>**median**<br>**cutoff**<br>**value**<br>**(0.1010**<br>**pg/mL),**<br>**n (%)** | RR (95% CI) | aRR (95% CI);<br>*CRI Model* | p-value;<br>*CRI Model* | aRR<br>(95% CI);<br>*Influenza*<br>*Model* | p-value;<br>*Influenza*<br>*Model* |
| --- | --- | --- | --- | --- | --- | --- | --- | --- | --- |
| CRI Status |  |  |  |  |  |  |  |  |  |
|  | Without<br>CRI | 61 | 31<br>(50.8) | 30 (49.2) | Ref. | Ref. |  |  |  |
|  | With CRI | 78 | 39<br>(50.0) | 39 (50.0) | 1.02 (0.73–<br>1.43) | 0.97 (0.72–<br>1.29) | 0.82 | - | - |
| Influenza<br>Status |  |  |  |  |  |  |  |  |  |
| Among<br>Cardiac<br>Group** |  |  |  |  |  |  |  |  |  |
|  | Negative | 91 | 33<br>(36.3) | 58 (63.7) | Ref. | - | - | Ref. |  |
|  | Positive | 9 | 3<br>(33.3) | 6 (66.7) | 1.12 (0.20–<br>2.44) | - | - | 1.02<br>(0.44–<br>2.37) | 0.97 |
| Among<br>Healthy<br>Group** |  |  |  |  |  |  |  |  |  |
|  | Negative | 36 | 33<br>(91.7) | 3 (8.3) | Ref. | - | - | Ref. |  |
|  | Positive | 3 | 1<br>(33.3) | 2 (66.7) | 13.6 (1.39–<br>133.40) | - | - | 8.18<br>(1.34–<br>49.82) | 0.02 |
| Age* |  |  |  |  |  |  |  |  |  |
|  | 40–64<br>years | 112 | 59<br>(52.7) | 53 (47.3) | Ref. | Ref. |  | Ref. |  |
|  | >65<br>years | 27 | 11<br>(40.7) | 16 (59.3) | 1.25 (0.87–<br>1.81) | 1.04 (0.75–<br>1.45) | 0.8 | 1.01<br>(0.57–<br>1.81) | 0.97 |
| Gender* |  |  |  |  |  |  |  |  |  |
|  | Male | 134 | 67<br>(50.0) | 67 (50.0) | Ref. | Ref. |  | Ref. |  |
|  | Female | 5 | 3<br>(60.0) | 2 (40.0) | 0.80 (0.27–<br>2.37) | 0.72 (0.25–<br>2.02) | 0.53 | 0.74<br>(0.18–<br>3.07) | 0.68 |
| Past Medical<br>History* |  |  |  |  |  |  |  |  |  |
|  | No | 56 | 32<br>(57.1) | 24 (42.9) | Ref. | Ref. |  | Ref. |  |
|  | Yes | 83 | 38<br>(45.8) | 45 (54.2) | 1.27 (0.88–<br>1.82) | 1.12 (0.82–<br>1.53) | 0.49 | 1.15<br>(0.69–<br>1.91) | 0.6 |
| History of<br>Tobacco Use* |  |  |  |  |  |  |  |  |  |
|  | No | 44 | 23<br>(52.3) | 21 (47.7) | Ref. | Ref. |  | Ref. |  |
|  | Yes | 95 | 47<br>(49.5) | 48 (50.5) | 1.06 (0.73–<br>1.53) | 0.93 (0.68–<br>1.25) | 0.62 | 1.00<br>(0.59–<br>1.69) | 1.00 |
| Physical<br>Activities* |  |  |  |  |  |  |  |  |  |
|  | No | 65 | 29<br>(44.6) | 36 (55.4) | Ref. |  | - | Ref. | - |
|  | Yes | 74 | 41<br>(55.4) | 33 (44.6) | 0.81 (0.58–<br>1.13) |  | - | 0.87<br>(0.54–<br>1.41) | 0.58 |

| Participant Groups* |  |  |  |  |  |  |  |  |  |
| --- | --- | --- | --- | --- | --- | --- | --- | --- | --- |
|  | Healthy group | 39 | 34 (87.2) | 5 (12.8) | Ref. | Ref. | <0.001 | - | - |
|  | Cardiac group | 100 | 36 (36.0) | 64 (64.0) | 5.00 (2.10–11.92) | 4.84 (2.11–11.16) | <0.001 | - | - |

**Note:**
\*Both the covariates for the CRI multivariable model and those for the influenza multivariable model are presented in the same table.
\*\*The association between influenza status and IL-6 levels is stratified by participant groups (Healthy vs. Cardiac), demonstrating effect modification where the association was significant only among the healthy participant group.

## 4. DISCUSSION

In this study, participants with recent CRI or confirmed influenza had higher inflammatory biomarker blood levels, although differences were not significant. However, influenza was significantly associated with high IL-6 levels only in healthy participants, indicating possible effect modification. Moreover, our analyses identified significant links between acute cardiac illnesses and blood levels of these high-risk markers. Overall, our results do not support the blood levels of these biomarkers as reliable predictors of imminent acute cardiac events, specifically following an acute CRI or influenza episode. Nevertheless, elevated biomarker levels may indicate inflammation that could exacerbate underlying cardiovascular conditions, regardless of CRI or influenza history. Further studies with larger sample sizes and more robust designs are needed to better elucidate the impact of recent respiratory infections on biomarker dynamics and their potential role as early indicators of imminent acute cardiac events.

No epidemiological studies have directly examined how cytokine blood levels change after respiratory illnesses like influenza by comparing affected and unaffected individuals and relating these changes to the risk of acute cardiac events, revealing a significant gap in research. Existing clinical research primarily targets patients with severe respiratory infections, exploring how elevated pro-inflammatory biomarkers relate to serious outcomes such as hospitalization or death, rather than to immediate cardiac risks (22, 37). Some recent studies have delved into the role of inflammatory markers in predicting different outcomes for influenza versus SARS-CoV-2, and how specific influenza strains may subtly alter cardiovascular functions. Studies have also investigated the potential for influenza A to trigger cytokine storms, the genetic factors influencing cytokine response to severe influenza, and how influenza may enhance cytokine production, thereby increasing systemic inflammation and complicating conditions like encephalopathy (38–46).

CRP and IL-6 are established biomarkers for severe respiratory infections, but their short-term role in predicting imminent cardiac events post-respiratory illness remains unexplored. This underscores the need to study the links between mild respiratory infections, cytokine changes, and cardiovascular risk. Our study uniquely investigates these associations in acute heart conditions, differing from other study designs (12, 47, 48). Variations in hs-CRP and IL-6 levels across studies arise from differences in design, participant traits, exclusion criteria, and sampling times. High variability indicates the complex blood dynamics of these biomarkers, suggesting that they may possess highly sensitive characteristics to various stimuli. This sensitivity necessitates individualized interpretation and renders them unreliable as standalone screening tools for early cardiac conditions due to their hyperresponsiveness to genetic, environmental, and lifestyle factors, comorbidities, and the timing of measurement (49–51). Consequently, standardized cutoffs may not accurately reflect an individual’s risk. Further research is essential to understand these discrepancies and their clinical implications.

During influenza seasons, respiratory infections may heighten AMI and stroke risks through immune imbalances, inflammatory markers IL-6 and CRP and plaque rupture (54–57). A recent large-scale RCT found influenza vaccination reduces adverse cardiovascular events, including hospitalization or death due to AMI, by about 40% (24). Vaccines may stabilize atherosclerotic plaques, suppress pro-inflammatory mediators, and stimulate anti-inflammatory responses, potentially preventing acute cardiac events like heart failure (25, 26). Personalized medicine using inflammatory markers and genetics could improve early detection and prevention of cardiovascular events in at-risk individuals, in alignment with clinical guidelines such as those by ACC and ESC (58–60).

Acute phase reactants and cytokines like hs-CRP and IL-6 can detect early inflammation but lack specificity for predicting imminent cardiac events after respiratory infections such as influenza. Challenges include non-specific indicators, absence of standardized measurements, and limited clinical utility. Consequently, inflammatory cytokine tests for acute cardiac events remain restricted to specific assays (61, 62). Monitoring cytokines like hs-CRP and IL-6 alongside conventional diagnostics like ECG may improve early detection of acute cardiac events during respiratory infections. However, their low specificity requires careful interpretation for accurate, cost-effective risk identification and treatment (63–65). Nevertheless, biomarkers like CRP are more reliable indicators of inflammation than IL-6 in sub-clinical coronary artery conditions (33, 52, 53).

### Strengths and Limitations

Our study is the first conducted in a low-income setting like Bangladesh to examine the relationship of blood levels of inflammatory biomarkers with both acute respiratory illnesses and cardiac events. A key strength was the inclusion of diverse study groups, including cardiac patients and healthy individuals, further classified based on recent acute respiratory illnesses or laboratory-confirmed influenza status. By simultaneously investigating cardiac conditions and respiratory illnesses within the same population, the study provided valuable insights into how high-risk inflammatory mediators in the blood relate to recent respiratory infections and subsequent cardiac events. We focused on high-sensitivity hs-CRP and IL-6, well-known pro-inflammatory mediators, as well as high cardiac risk markers.

However, our study has several notable limitations. The descriptive cross-sectional design, constrained by limited resources, prevents establishing causal relationships. Additionally, the small sample size restricts the generalizability of our findings, as the study was intended to provide preliminary insights. Another limitation is the inability to confirm respiratory infections among all the participants reporting recent CRI through comprehensive laboratory tests. This, coupled with the subjectivity and limited specificity of CRI definitions, may not exclude some misclassifications.

Furthermore, the healthy participant group did not undergo cardiac assessments like ECG or ETT to exclude subclinical coronary artery diseases, especially among those with cardiovascular risk factors.

In the current study, blood samples for cytokine assessment were collected only at baseline, which may not have consistently coincided with peak cytokine elevations following respiratory illness onset. The lack of a standardized window period meant that some samples were taken during peak cytokine levels while others were taken during the declining phase. The dynamic fluctuations of cytokines (19) present practical challenges for their use as early inflammatory cardiac markers, as rapid changes can result in inaccurate measurements and potential false diagnoses. In chronic conditions, sustained elevated cytokine levels complicate distinguishing between normal background levels and significant rises due to an acute respiratory event. In summary, while our study offers preliminary insights into the role of inflammatory biomarkers in the context of recent respiratory infections and acute cardiac events, further research is essential.

## 5. CONCLUSION

The current study’s findings do not clarify the significance of inflammatory biomarkers like hs-CRP and IL-6 in assessing the short-term risk of acute cardiac events following recent respiratory illnesses. Currently, these biomarkers cannot be broadly recommended for early detection of events such as AMI after respiratory episodes due to specificity issues and complex circulatory dynamics. Further research is necessary to determine their clinical role more precisely. Future studies should adopt more robust study designs, such as large sample size and longitudinal cohort studies, to better examine these complex interactions. While past studies have focused on using biomarkers to confirm acute cardiac events, there is a growing need for reliable markers for early risk stratification, and despite their current limitations, inflammation marker assays may play an increasingly important role in future research for the early detection of adverse cardiac events.

## Acknowledgements

This researchstudy was funded by Swedish International Development Cooperation Agency (Sida). This supporting source was not involved in the study design, collection, analysis, and interpretation of data, writing of the report, and the decision to submit the report for publication. icddr,b acknowledges with gratitude the commitment of Sida to its research efforts. icddr,b is also grateful to the Governments of Bangladesh, and Canadaor providing core/unrestricted support. The authors are also grateful to the study data collection team and study participants for their valuable data. The authors also acknowledge the support of NICVD authority in conducting this study.

## Author contribution

**M.A.A:** Conceptualization, Funding acquisition, Methodology, Investigation, Project administration, Data curation, Visualization, Formal analysis, Writing- original draft, Writing- review & editing, Approval of final manuscript. **C.R.M:** Conceptualization, Methodology, Investigation, Project administration, Resources, Writing- review & editing, Approval of final manuscript. **B.R**: Investigation, Formal analysis, Writing- review & editing, Approval of final manuscript. **M.Z.R**: Investigation, Resources, Writing- review & editing, Approval of final manuscript. **M.R**: Investigation, Resources, Writing-review & editing, Approval of final manuscript. **A.K.M.M.I**: Conceptualization, Methodology, Investigation, Project administration, Resources, Writing- review & editing, Approval of final manuscript.

**P.K.G**: Visualization, Formal analysis, Writing- review & editing, Approval of final manuscript. **Z.A**: Investigation, Formal analysis, Writing- review & editing, Approval of final manuscript. **F.C**: Investigation, Formal analysis, Writing- review & editing, Approval of final manuscript. **F.Q**: Conceptualization, Methodology, Investigation, Project administration, Resources, Writing- review & editing, Approval of final manuscript. **A.A.C**: Supervision, Methodology, Formal analysis, Writing- review & editing, Project administration, Approval of final manuscript.

## Ethical standard

The authors assert that all procedures contributing to this work comply with the ethical standards of the relevant national and institutional committees on human experimentation and with the Helsinki Declaration of 1975, as revised in 2008. Enrolment of the participants started after approval of the study by the ICDDR,B Institutional Review Board (PR-17039) and UNSW Human Research Ethics Committee (HC 17861). Informed written consent to participate in the study was obtained.

## Data availability statement

Data generated during the study are subject to a data access policy of icddr,b and are available from icddr,b’s research administration on reasonable request through the corresponding author. No additional data are available. The corresponding author, has full access to all of the data in this study and takes complete responsibility for the integrity of the data and the accuracy of the data analysis.

## Patient and public involvement

Patients and/or the public were not involved in the design, conduct, reporting, or dissemination plans of this research.

## Provenance and peer review

Not commissioned; externally peer-reviewed.

## Transparency statement

The corresponding author affirms that this manuscript is an honest, accurate, and transparent account of the study being reported; that no important aspects of the study have been omitted; and that any discrepancies from the study as planned (and, if relevant, registered) have been explained.

